# Adherence to National Standard Treatment Guidelines in the Management of Hypertension and Associated Factors Among Healthcare Providers at Public District Hospitals in Dar es Salaam, Tanzania

**DOI:** 10.64898/2025.12.29.25343177

**Authors:** Gasper Singfrid Mung’ong’o, Daniel Msilanga, Paul Alikado Sabuni, Humphrey Godwin Medarakini, Gladys Mahiti, Rose Mpembeni

**Affiliations:** Department of Development Studies, Muhimbili University of Health and Allied Sciences, P.O. Box 65001, Dar es Salaam, Tanzania; Department of Epidemiology and Biostatistics, Muhimbili University of Health and Allied Sciences, P.O. Box 65001, Dar es Salaam, Tanzania; Department of Internal Medicine, Muhimbili University of Health and Allied Sciences, P.O. Box 65001, Dar es Salaam, Tanzania; Department of Epidemiology and Biostatistics, Catholic University of Health and Allied Sciences, P.O. Box 1464, Mwanza, Tanzania; Department of Emergency Medicine, Muhimbili University of Health and Allied Sciences, P.O. Box 65001, Dar es Salaam, Tanzania

## Abstract

**Background:** Hypertension affects 15–25% of adults in Tanzania, with less than 30% of treated patients achieving adequate blood pressure control, leading to complications like stroke, heart failure, and renal disease. The National Standard Treatment Guidelines (NSTGs), 6th edition (2021), guide management, but healthcare provider adherence in district hospitals is unclear.

**Materials and Methods:** A mixed-methods cross-sectional study was conducted at five public district hospitals in Dar Es Salaam, Tanzania. Quantitative data from 397 patient files assessed healthcare provider adherence in pharmacological treatment, complication monitoring, and comorbidity screening, categorized as complete, partial, or non-adherent. Modified Poisson regression was used to identify patient and provider factors. Qualitative data from 11 in-depth interviews were thematically analyzed for barriers, and facilitators to this adherence

**Results:** Only 26.2% of patients received management with complete adherence to NSTGs, 27.7% showed partial adherence, and 46.1% were non-adherent. Higher healthcare provider adherence was associated with follow-up visits (aPR = 5.81; 95% CI: 3.12–10.81), Grade 1 hypertension (aPR = 3.87; 95% CI: 1.95–7.65), Grade 2 hypertension (aPR = 5.06; 95% CI: 2.64–9.71), presence of comorbidities (aPR = 2.35; 95% CI: 1.31–4.22), specialist providers (aPR = 7.00; 95% CI: 3.08–15.92), and provider experience of 5–10 years (aPR = 3.25; 95% CI: 1.52–6.97) and > 10 years (aPR = 3.91; 95% CI: 1.88–8.14). Barriers included outdated guidelines, absence of comorbidity-specific protocols, limited user input, weak monitoring, drug shortages, inadequate diagnostics, staffing gaps, and inadequate training, while facilitators included peer collaboration, mentorship, accessible guidelines, and continuing medical education.

**Conclusion:** Low healthcare provider adherence to NSTGs contributes to poor hypertension control. Interventions like updated and inclusive guidelines, enhanced training, improved resource availability, and peer support are critical to improve adherence and patient outcomes.

## Introduction

Hypertension remains a major global public health challenge and one of the leading causes of mortality and disability worldwide. The number of people living with hypertension has doubled from an estimated 650 million in 1990 to 1.3 billion by 2019. (1,2) This increase is strongly associated with modifiable risk factors such as obesity, sedentary lifestyles, and unhealthy dietary patterns, particularly high salt and fat consumption (3–5). Urbanization has further accelerated this trend, with rapid economic and social transitions contributing to reduced physical activity and higher exposure to behavioral risk factors (5–7).

Globally, hypertension is responsible for approximately 10.8 million deaths annually and accounts for an estimated 235 million disability-adjusted life years (DALYs) lost. The health risks associated with hypertension include stroke, myocardial infarction, renal impairment, and premature death. (2,8,9).

Non-Communicable Diseases (NCDs), including hypertension, have been prioritized within the Sustainable Development Goal (SDG) 3.4, which aims to reduce premature mortality from NCDs by one-third by 2030.(10) Despite global efforts, only about 23% of individuals with hypertension achieve recommended control levels through treatment.(11) Control rates in Sub-Saharan Africa are among the lowest globally, largely due to issues such as poor treatment adherence, limited medication availability, and insufficient healthcare follow-up systems. (12)

In Tanzania, the burden of NCDs has grown steadily over the past decades. NCDs accounted for 19% of total DALYs in 1990, rising to 34% by 2015, and currently contribute to nearly one-third of all deaths, with a substantial proportion occurring prematurely between ages 30 and 70. (13,14)

The prevalence of hypertension in Tanzania is particularly high in urban areas, reaching up to 23.7%, compared with about 17.6% in rural populations (13,14). Additionally, a large proportion of Tanzanians with hypertension remain undiagnosed due to limited screening and inadequate public health education, especially in rural communities (14–16). However, among the patients who are on treatment less than 30% have achieved the target blood pressure placing them at risk for the life threatening complications and premature mortality. (14,17–20)

Recognizing these trends, Tanzania’s National Strategic Plan for the Prevention and Control of Non-Communicable Diseases (2021–2026) identifies hypertension control as a national priority and aligns with both the Health Sector Strategic Plan V and the Universal Health Coverage agenda. A key strategy within this framework is the standardization of care given to Hypertensive patients. (21)

The National Standard Treatment Guidelines (NSTGs), developed by the Ministry of Health since 1991 and most recently revised in 2021, provide evidence-based guidance for the diagnosis and management of hypertension in Tanzania. (22,23). However the extent of healthcare provider adherence to the NSTGs in management of Hypertension remains unclear.

Assessing healthcare provider adherence to the NSTGs and the factors influencing it is therefore essential for improving hypertension care in Tanzania. This study aims to evaluate adherence to the guidelines among healthcare providers in public district hospitals in Dar es Salaam and to identify patient-, provider-, and health system–related factors associated with adherence. The findings will inform targeted interventions to strengthen guideline compliance, enhance hypertension management, and ultimately improve patient outcomes.

## Materials and Methods

### Study design

An analytical cross-sectional design using a mixed-methods approach was used where the quantitative component assessed the level of healthcare provider adherence to NSTGs and identified statistically associated provider and patient factors, while the qualitative component explored barriers and facilitators that influence this adherence.

### Study Area

The study was carried out at five public district hospitals in Dar Es Salaam region one from each district. These are: Kivule district hospital (Ilala), Ubungo district hospital (Ubungo), Mbagala rangi tatu hospital (Temeke), Kigamboni district hospital (Kigamboni) and Mabwepande district hospital (Kinondoni). These hospitals are located in Dar Es Salaam, an urban setting where there is a growing Hypertensive patient population and hence an appropriate setting to evaluate healthcare provider adherence to the NSTGs in management of these patients. (26)

### Study Population

The study population included patients diagnosed with hypertension who attended outpatient departments at the study facilities between 1 January and 31 December 2024, and healthcare providers of all cadres involved in prescribing and managing hypertension. Quantitative data were extracted from patient records, while qualitative data were collected through in-depth interviews with healthcare providers. Both quantitative data extraction and qualitative interviews were conducted at the study facilities between 1 April and 31 May 2025.

### Inclusion and Exclusion Criteria

#### 1. Inclusion Criteria

i. All patients with Hypertension who were attended from January 2024 to December 2024 at the selected public district hospitals and have case file.
ii. All healthcare providers in the public district hospitals who prescribe management for Hypertensive patients

#### 2. Exclusion criteria

i. All patients with case files that do not have a documented diagnosis and management provided between January 2024 and December 2024.
ii. All healthcare providers with less than one year of experience working in the selected hospitals as they have limited exposure to the Tanzanian health system and the NSTG.

### Sample Size and Sampling Procedures

For the quantitative analysis, all eligible hypertensive patient records from January to December 2024 were included to minimize selection bias. Duplicate entries were identified through unique Medical Record Numbers and removed to ensure each patient contributed only once to the dataset.

For the qualitative component, eleven healthcare providers participated in in-depth interviews. Sampling followed the principle of data saturation. After the ninth interview, no new themes emerged, and two additional interviews were conducted to confirm thematic stability. By the eleventh interview, redundancy was reached, indicating sufficient depth and breadth of data across provider cadres and facility contexts.

Patient data were extracted from the Government of Tanzania Hospital Management Information System (GOTHOMIS) outpatient department registers. Providers were selected purposively based on their direct involvement in hypertension management at either Non-Communicable Disease (NCD) clinics or general outpatient departments. Only those with at least one year of service and who gave written informed consent to participate were included.

### Study variables

The dependent variable was healthcare provider adherence to NSTGs in hypertension management, measured through a structured checklist applied to retrospective patient records covering the three months preceding the most recent clinic visit.

Adherence was assessed in three domains:

1. **Pharmacological management:** Consistency of prescribed antihypertensive medications with NSTG recommendations based on patient classification, comorbidities, and crisis status.
2. **Monitoring for complications:** Performance of recommended investigations such as ECG, echocardiography, renal function tests, urinalysis, fundoscopy, and relevant laboratory tests.
3. **Screening for comorbidities:** Screening for diabetes mellitus, dyslipidemia, and obesity through glucose tests, lipid profiles, and BMI assessment.

Each item was scored dichotomously (1 = adherent, 0 = non-adherent). The adherence score was computed as a percentage of total possible points. Independent variables included patient characteristics (age, sex, BMI, payment mode, visit type, hypertension grade, comorbidities) and provider characteristics (age, sex, professional cadre, years of experience, and facility). The qualitative component explored barriers and facilitators influencing provider adherence to NSTGs.

### Quantitative Data collection

Data were collected by the principal investigator using a structured digital checklist developed on the ODK platform. Eligible hypertensive patient records were identified via GOTHOMIS registers and verified through NCD clinic and laboratory registers. The checklist captured patient demographics, clinical characteristics, investigations, and treatment in line with NSTG recommendations. Data were entered in real time into the ODK platform and securely uploaded to a central server for analysis.

### Qualitative Data collection

In-depth interviews were conducted in Swahili by the principal investigator using a structured guide. All interviews were audio-recorded and supplemented with detailed field notes capturing non-verbal cues and contextual observations. Recordings were transcribed verbatim and translated into English by trained bilingual research assistants.

### Data Management

Audio recordings were stored securely on password-protected Google Drive folders accessible only to the investigator. Transcripts were anonymized before translation. Quantitative data extracted from ODK were exported to Microsoft Excel and stored in encrypted drives before analysis in Stata version 17. Regular backups were maintained to ensure data security and integrity.

### Validity and Reliability

The quantitative data collection tool was developed based on the 6th Edition NSTGs and reviewed by two clinical experts (a nephrologist and an emergency physician) for content validity. The tool was pretested on 50 hypertensive patient files at a private outpatient clinic to assess clarity and usability, leading to minor revisions. Face validity was confirmed by three independent medical practitioners who reviewed the final checklist for relevance and comprehensiveness.

### Trustworthiness

Trustworthiness was ensured through the application of four key criteria. Credibility was enhanced by prolonged engagement of principal investigator in the field, peer debriefing, and triangulation with observational data to ensure accurate representation of participants’ views. Transferability was supported through rich, detailed descriptions of the study context and participant selection, enabling readers to assess the applicability of the findings to other settings. Dependability was maintained by establishing an audit trail that documented all methodological steps, ensuring consistency and replicability of the research process. Finally, confirmability was achieved through peer debriefing and member checking, which helped minimize researcher bias and ensure that the findings accurately reflected participants’ perspectives.

### Data analysis

Data were analyzed using Stata version 17. Descriptive statistics summarized participant characteristics: means and standard deviations for normally distributed variables, medians and interquartile ranges for skewed data, and frequencies and proportions for categorical variables.

Adherence scores were calculated using:

Adherence score= (Points scored / Total points available) x 100%

Scores were categorized as complete (≥65%), partial (50–64.9%), or non-adherent (<50%), consistent with prior regional studies. (27,28).

Bivariate analyses were conducted to identify variables significantly associated with adherence (p < 0.05). These variables, along with age and sex, were included in the multivariable model. Statistical significance was defined as p < 0.05.

### Qualitative data analysis

Thematic analysis was conducted using ATLAS.ti version 22. Transcripts were read repeatedly to ensure familiarity with the data. Relevant statements were coded inductively, grouped into subthemes, and then aggregated into major themes representing barriers and facilitators to adherence. Coding and theme development were iterative, with continuous comparison to ensure internal coherence and alignment with study objectives.

### Ethical Issues

Ethical approval was obtained from the Muhimbili University of Health and Allied Sciences (MUHAS) Research and Publications Committee (approval number DA.282/298/01.C/2694) on 11 March 2025. Administrative permission was granted by the Regional Medical Officer, District Medical Officers, and facility leadership. Written informed consent was obtained from all healthcare providers who participated in the in-depth interviews.

During data collection, the research team accessed patient medical records containing identifiable information. However, no personal identifiers were recorded or retained. Patient data were anonymized at the point of extraction, and individuals were identified only using unique medical record numbers for study purposes. Confidentiality was ensured through anonymization of data and secure data handling procedures.

## Results

### Patient characteristics

A total of 474 case files with unique medical record numbers of hypertensive patients attended at the public district hospitals from January 2024 to December 2024 were obtained from the GOTHOMIS OPD registers. Of these, 397 case files (83.8%) contained complete information including diagnosis, treatment, and monitoring for complications and comorbidities.

**Figure 1:**
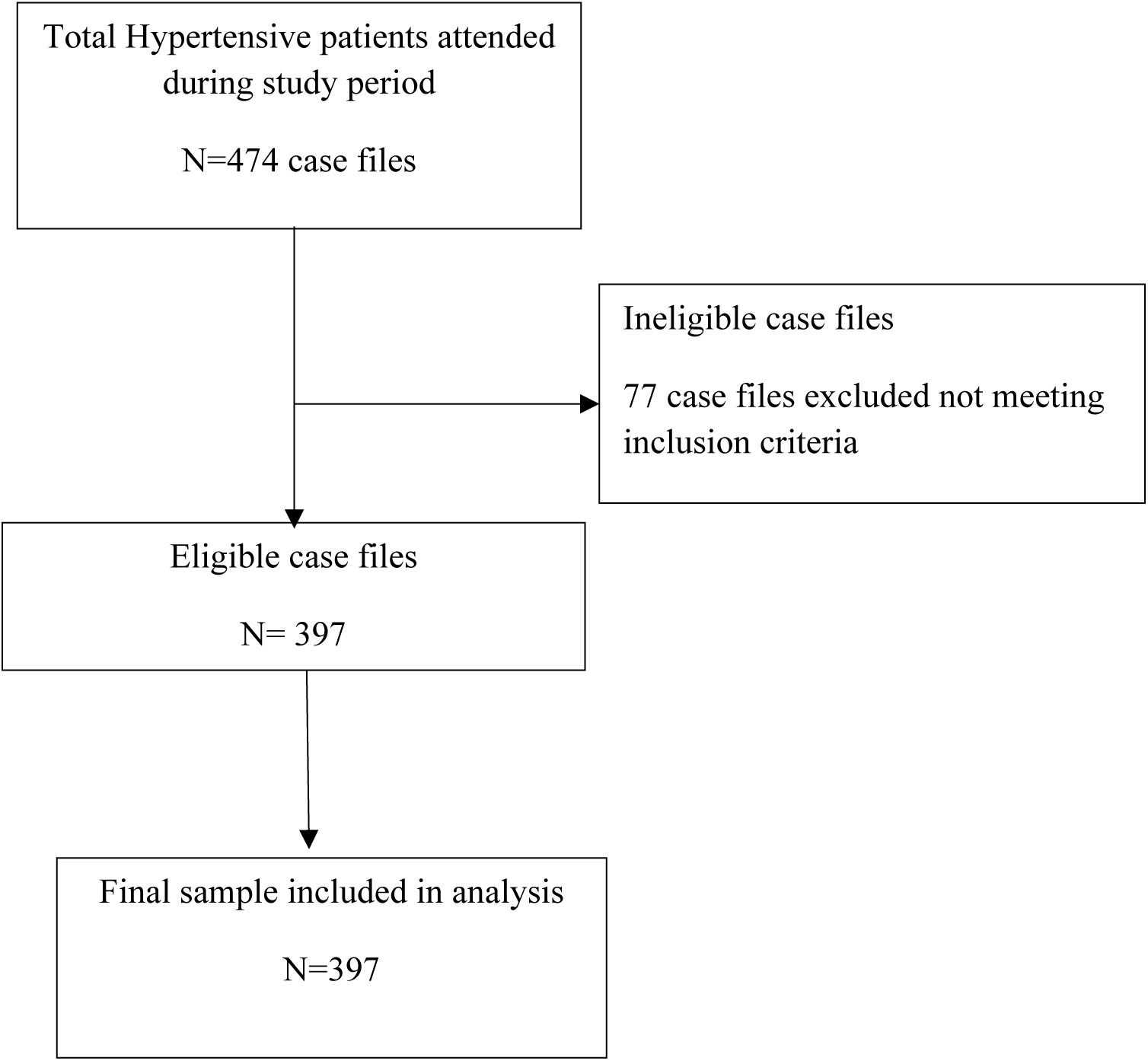
Enrollment of case files in the study

The median age of patients was 55 years (IQR: 52–61), with the majority (233; 58.7%) aged 45–60 years, and 39 patients (9.8%) aged below 45 years. Most patients were female (314; 79.1%). The mean body mass index (BMI) was 27.34 kg/m² (SD: 3.08), with 259 (65.2%) overweight and 81 (20.4%) obese. Regarding payment mode, 265 patients (66.8%) paid out-of-pocket, while 71 (17.9%) were covered by national health insurance. Follow-up visits accounted for 353 (88.9%) of encounters. Blood pressure control was achieved in 166 patients (41.8%), while 21 (5.3%) presented in hypertensive crisis. The median number of antihypertensive drugs used was 2 (range 1–4), with 268 patients (67.5%) using more than one drug. Comorbidities were present in 159 patients (40.0%), with obesity (20.4%) and diabetes mellitus (15.1%) being most common.

**Table 1a:** Demographic and clinical characteristics of the patients (n=397)

| Variable | Number | Percentage |
| --- | --- | --- |
| **Age (median, IQR)** | 55, 52-61 |  |
| **Age group** |  |  |
| Less than 45 | 39 | 9.8% |
| 45-60 | 232 | 58.4% |
| Above 60 | 126 | 31.7% |
| **Sex** |  |  |
| Female | 314 | 79.1% |
| Male | 83 | 20.9% |
| **Body mass index (mean, SD)** | 27.34, 3.083 |  |
| **Body mass index category** |  |  |
| Normal | 57 | 14.4% |
| Overweight | 259 | 65.2% |
| Obese | 81 | 20.4% |
| **Payment mode** |  |  |
| National health insurance | 71 | 17.9% |
| Government exception | 24 | 6.1% |
| iCHF | 37 | 9.3% |
| Cash | 265 | 66.8% |
| **Type of the visit** |  |  |
| New case | 44 | 11.1% |
| Follow up visit | 353 | 88.9% |
| **Grade of hypertension** |  |  |
| Controlled | 166 | 41.8% |
| Grade 1 | 188 | 47.4% |
| Grade 2 | 22 | 5.5% |
| Grade 3 | 21 | 5.3% |
| **Hypertensive crises** |  |  |
| Not in crisis | 376 | 94.7% |
| Hypertension emergency | 14 | 3.5% |
| Hypertension urgency | 7 | 1.8% |
| **Number of antihypertensives (median, range)** | 2, 1-4 |  |
| **Number of antihypertensives** |  |  |
| One | 129 | 32.5% |
| More than one | 268 | 67.5% |
| **Comorbidity condition** |  |  |
| No | 238 | 60.0% |
| Yes | 159 | 40.0% |
| **Types of comorbidity condition † (n=397)** |  |  |
| Diabetes | 60 | 15.1% |
| Heart failure | 35 | 8.8% |
| Pregnancy induced | 17 | 4.3% |
| Asthma | 21 | 5.3% |
| Arrhythmias | 9 | 2.3% |
| Dyslipidemias | 26 | 6.5% |
| Obesity | 81 | 20.4% |

### Healthcare provider characteristics

A total of 26 healthcare providers attended the Hypertensive patients included in the study (Table 1b). Health care providers were recruited in the five district hospitals where Kigamboni, Kivule, and Mabwepande each had 4 providers, while Mbagala Rangi Tatu and Ubungo each had 7. The mean age of providers was 36.25 years (SD: 8.45), with the majority aged between 21–30 years (n = 12). More providers were female (n = 15) than male. The most common professional cadre was medical doctor (n = 16), while specialists were few (n = 3). The mean duration of work experience was 6.69 years (SD: 3.61), with most providers having less than five years of experience (n = 12).

**Table 1b:**
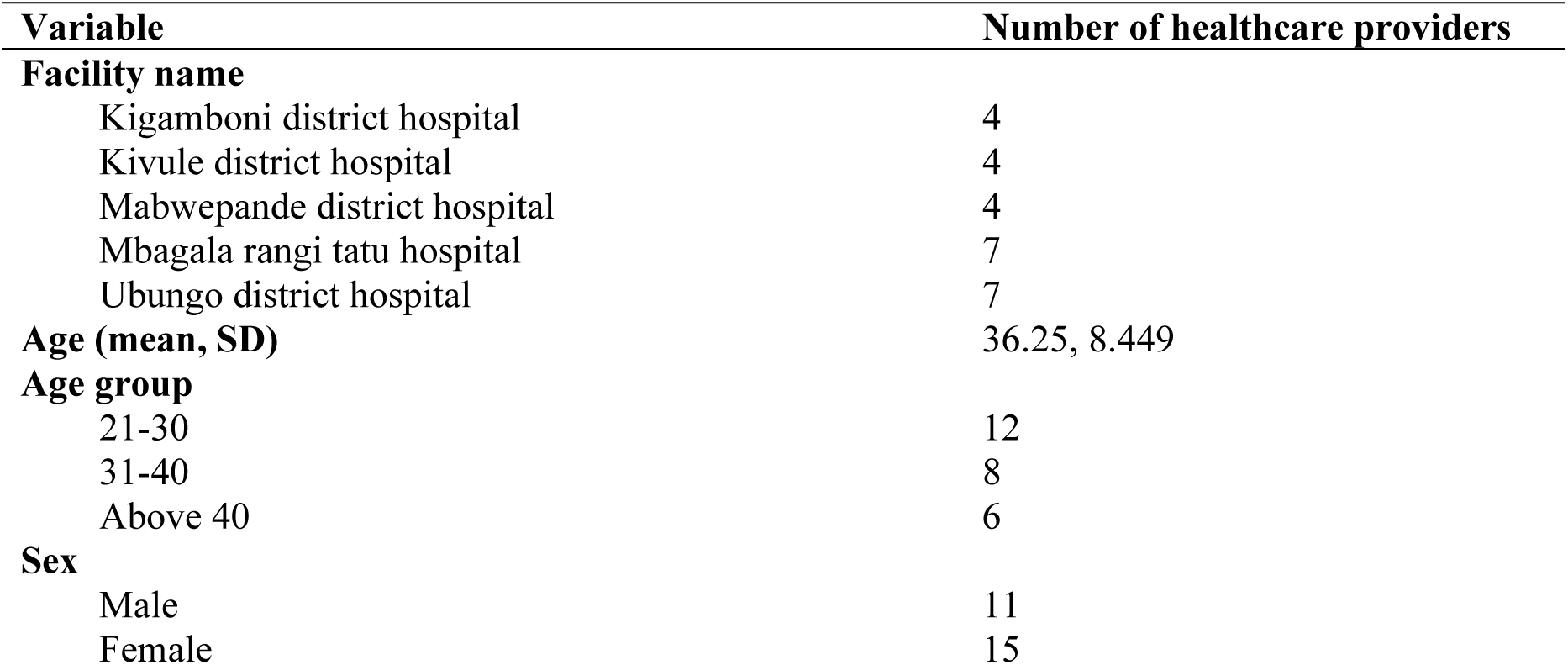
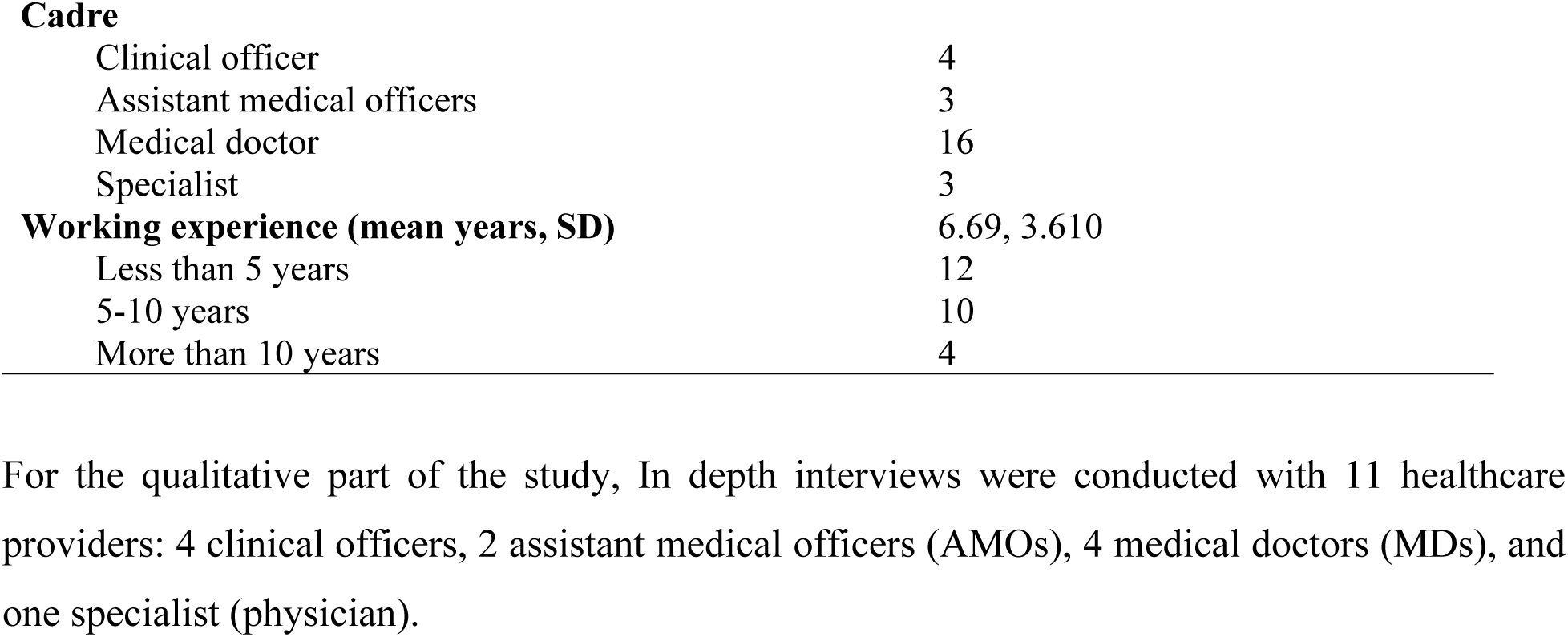
Demographic characteristics of the healthcare providers (n=26)

### Level of adherence of the healthcare providers

**Figure 2:**
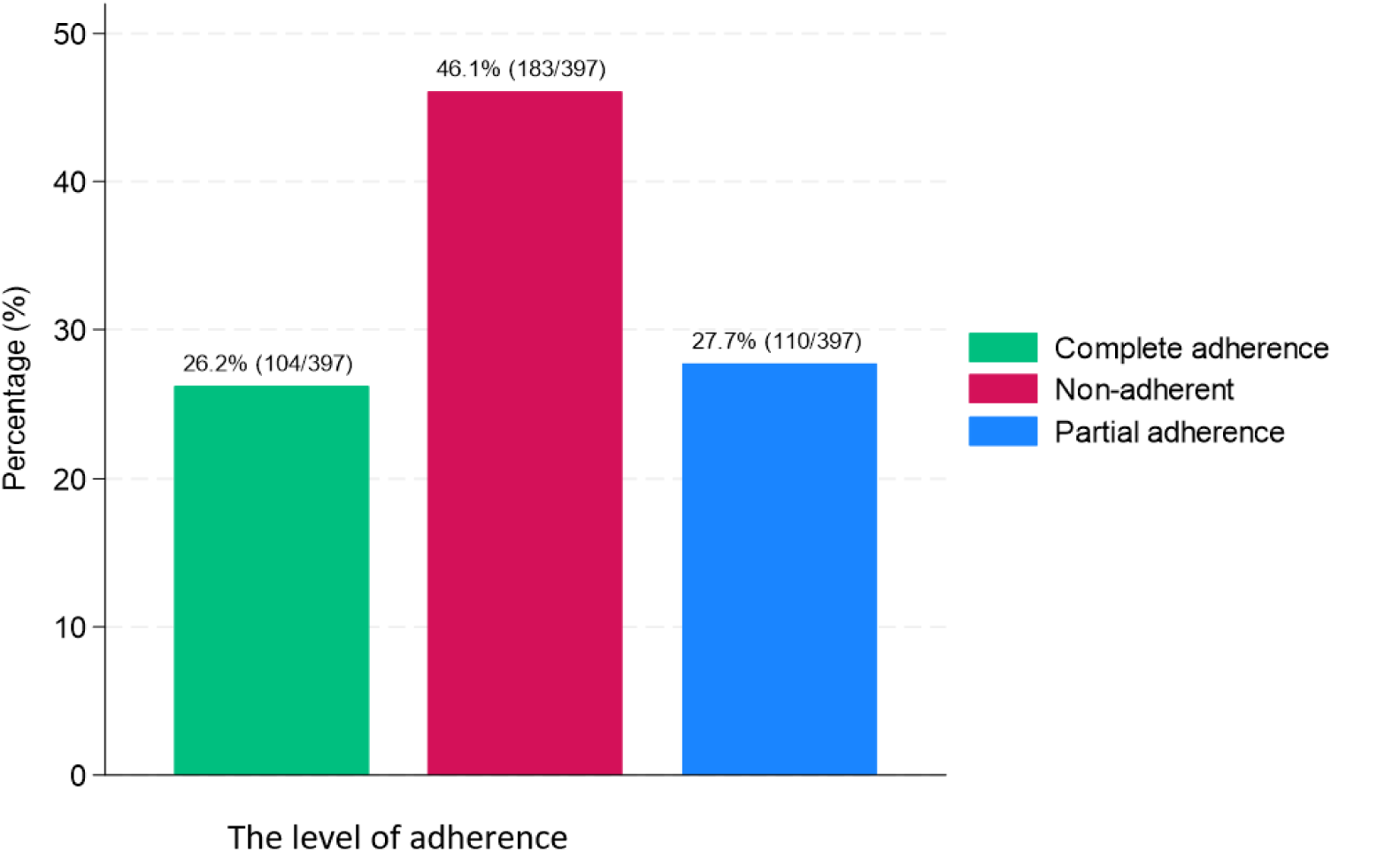
Healthcare provider adherence to NSTGs in management of Hypertension

### Patient factors associated with adherence of healthcare providers

Modified Poisson regression revealed that follow-up visits were significantly associated with guideline-concordant management compared to new cases (aPR = 5.81; 95% CI: 1.59–21.27; p = 0.008). Higher grades of hypertension were also positively associated with adherence, including Grade 1 (aPR = 3.87; 95% CI: 1.99–7.53) and Grade 2 (aPR = 5.06; 95% CI: 1.52–16.73). Presence of comorbidities doubled the likelihood of receiving guideline-adherent care (aPR = 2.35; 95% CI: 1.25–4.41; p = 0.008).

**Table 2:** Patient factors associated with adherence of healthcare providers to the NSTGs in the management of hypertension.

| Variable | Total | Adherence status | | Unadjusted PR $\delta$ | | Adjusted PR $\delta$ | |
| --- | --- | --- | --- | --- | --- | --- | --- |
|  |  | No<br>n (%) | Yes<br>n (%) | PR, 95% CI | p-value | PR, 95% CI | p-value |
| **Age group** |  |  |  |  |  |  |  |
| Less than 45 | 39 | 32(82.1) | 7(18.0) | 1 |  | 1 |  |
| 45-60 | 232 | 171(73.7) | 61(26.3) | 1.37, 0.53-3.53 | 0.515 | 0.84, 0.27-2.61 | 0.766 |
| Above 60 | 126 | 90(71.4) | 36(28.6) | 1.60, 0.59-4.29 | 0.348 | 0.59, 0.17-2.03 | 0.406 |
| **Sex** |  |  |  |  |  |  |  |
| Female | 314 | 231(73.6) | 83(26.4) | 1 |  | 1 |  |
| Male | 83 | 62(74.7) | 21(25.3) | 0.98, 0.53-1.81 | 0.952 | 1.47, 0.71-3.08 | 0.302 |
| **Body mass index category** |  |  |  |  |  |  |  |
| Normal | 57 | 40(70.2) | 17(29.8) | 1 |  |  |  |
| Overweight | 259 | 201(77.6) | 58(22.4) | 0.68, 0.33-1.39 | 0.289 |  |  |
| Obese | 81 | 52(64.2) | 29(35.8) | 1.26, 0.55-2.88 | 0.577 |  |  |
| **Payment mode** |  |  |  |  |  |  |  |
| Cash | 265 | 204(77.0) | 61(23.0) | 1 |  |  |  |
| Insurance | 132 | 89(67.4) | 43(32.6) | 1.31, 0.77-2.23 | 0.315 |  |  |
| **Type of the visit** |  |  |  |  |  |  |  |
| New case | 44 | 40(90.9) | 4(9.1) | 1 |  | 1 |  |
| Follow up visit | 353 | 253(71.7) | 100(28.3) | 3.23, 1.07-9.78 | **0.038** | 5.81, 1.59-21.27 | **0.008** |
| **Grade of hypertension** |  |  |  |  |  |  |  |
| Controlled | 166 | 139(83.7) | 27(16.3) | 1 |  | 1 |  |
| Grade 1 | 188 | 131(69.7) | 57(30.3) | 3.21, 1.77-5.84 | **<0.001** | 3.87, 1.99-7.53 | **<0.001** |
| Grade 2 | 22 | 12(54.6) | 10(45.5) | 5.27, 1.81-15.33 | **0.002** | 5.06, 1.52-16.73 | **0.008** |
| Grade 3 | 21 | 11(52.4) | 10(47.6) | 3.88, 1.33-11.30 | **0.013** | 2.78, 0.76-10.13 | 0.122 |
| **Comorbidity condition** |  |  |  |  |  |  |  |
| No | 238 | 194(81.5) | 44(18.5) | 1 |  | 1 |  |
| Yes | 159 | 99(62.3) | 60(37.7) | 2.98, 1.75-5.08 | **<0.001** | 2.35, 1.25-4.41 | **0.008** |

Hints: PR=Prevalence Ratio; 95% CI=95 percent confidence interval; $\delta$ =Adjusted by cluster i.e., healthcare providers; $\sigma_u=0.993$ ; $\rho=0.23$ , $\chi^2=30.95$ ( $p<0.001$ )

### Healthcare providers’ factors associated with adherence

Provider cadre and experience influenced adherence. Specialists demonstrated significantly higher adherence compared to clinical officers (aPR = 7.00; 95% CI: 1.05–46.55; p = 0.004). Providers with 5–10 years of experience (aPR = 3.25; 95% CI: 1.17–9.03; p = 0.024) and more than 10 years of experience (aPR = 3.91; 95% CI: 1.20–12.73; p = 0.023) were also more likely to adhere. Age and sex showed no significant association after adjustment. **Table 3** presents provider-related factors associated with adherence.

**Table 3:** Healthcare providers’ factors associated with adherence of healthcare providers to the NSTGs in the management of hypertension (n=397)

| Variable | Total patient attended | Adherence status | | Unadjusted PR $\delta$ | | Adjusted PR $\delta$ | |
| --- | --- | --- | --- | --- | --- | --- | --- |
|  |  | No n (%) | Yes n (%) | PR, 95% CI | p-value | PR, 95% CI | p-value |
| **Age group** |  |  |  |  |  |  |  |
| 21-30 | 140 | 120(85.7) | 20(14.3) | 1 |  | 1 |  |
| 31-40 | 158 | 100(63.3) | 58(36.7) | 2.92, 1.08-7.86 | **0.034** | 0.48, 0.15-1.52 | 0.214 |
| Above 40 | 99 | 73(73.7) | 26(26.3) | 2.21, 0.76-6.51 | 0.147 | 0.86, 0.30-2.47 | 0.783 |
| **Sex** |  |  |  |  |  |  |  |
| Male | 160 | 117(73.1) | 43(26.9) | 1 |  | 1 |  |
| Female | 237 | 176(74.3) | 61(25.7) | 0.79, 0.29-2.07 | 0.624 | 1.28, 0.70-2.31 | 0.422 |
| **Cadre** |  |  |  |  |  |  |  |
| Clinical officer | 23 | 21(91.3) | 2(8.7) | 1 |  | 1 |  |
| Assistant medical officers | 39 | 34(87.2) | 5(12.8) | 1.56, 0.25-9.76 | 0.637 | 0.64, 0.09-4.27 | 0.645 |
| Medical doctor | 237 | 191(80.6) | 46(19.4) | 2.45, 0.51-11.69 | 0.262 | 1.48, 0.31-7.04 | 0.624 |
| Specialist | 98 | 47(48.0) | 51(52.0) | 11.66, 2.31-58.99 | **0.002** | 7.00, 1.05-46.55 | **0.004** |
| **Working experience (mean years, SD)** |  |  |  |  |  |  |  |
| Less than 5 years | 144 | 128(88.9) | 16(11.1) | 1 |  | 1 |  |
| 5-10 years | 177 | 117(66.1) | 60(33.9) | 3.45, 1.41-8.37 | **0.006** | 3.25, 1.17-9.03 | **0.024** |
| More than 10 years | 76 | 48(63.2) | 28(36.8) | 4.70, 1.61-13.73 | **0.005** | 3.91, 1.20-12.73 | **0.023** |

*Hints: PR=Prevalence Ratio; 95% CI=95 percent confidence interval; $\delta$ =Adjusted by cluster i.e.,* *patient attended*

### Barriers That Influence Healthcare Provider Adherence

Several barriers affecting healthcare provider adherence to the NSTGs in the management of Hypertension emerged from the interviews. These were organized into four main themes:

### Guideline-Related Barriers

The providers expressed concerns about the outdated nature of the NSTGs in the management of Hypertension, which have not been revised since 2021 and do not include newer, more effective medications.

> “Some things in the STG are outdated. Many new drugs exist but aren’t in the STG. A patient comes with a drug, you check the STG, it’s not there, yet it’s effective” (Medical Doctor).

Another provider highlighted the role of local research to support timely updates.

> “The Ministry could take the role to empower so that research is done here in our African, Tanzanian community….so that when they sit in meetings to update the STG, they can incorporate the findings. Also, they should update the STG, waiting 4/5 years is too long. At least 2 to 3 years, they should update earlier” (Specialist Physician).

Providers also noted the limited guidance provided by the NSTGs in the management of Hypertension for patients with comorbidities.

> “The management is generalized. For hypertension and diabetes, they’ve managed to explain which drugs to give or avoid, but for other categories, they haven’t explained well—the steps” (Medical Doctor).

Another provider highlighted the importance of involving frontline users in guideline development.

> “Also, STG development should involve users more; they should consult STG users. I don’t know if it’s cost or time. They should start with lower-level hospitals. Some things written in the STG, district hospitals can’t achieve due to limited resources and existing conditions” (Medical Doctor).

Policy restrictions were also identified as a barrier. A provider explained,

> “Some drugs are grade D or S, requiring a physician to prescribe… for NHIF patients, a Medical Officer can’t prescribe these drugs… the patients need to buy them. The only issue I see is when a patient has already been started on medication by a specialist, then comes here and finds me, an MD, who can’t prescribe at that level” (Medical Doctor).

Another provider emphasized the need for supervision,

> “If the guideline wants us to do something, they should come regularly to do supervision with us so they know if we are following the guideline. Yes, they come from the regional level. They take reports, see how many patients we’ve treated, how we’ve treated them. But when they come, they only go through the NCD clinic, they don’t go to every doctor” (Medical Doctor).

### Resource Barriers

Providers frequently reported unavailability of NSTG-recommended medications, especially for hypertensive emergencies. One provider described,

> “IV Medications, honestly, if you get them, it’s by luck. Hydralazine, you might get it by luck. Labetalol, there’s none. All these other IVs, there’s none. You have to mix and match. For example, a patient was using Losartan H, went to a cardiologist, was prescribed Telmisartan, but at our hospital, even though it’s a district hospital, it’s not available” (Assistant Medical Officer).

Insufficient diagnostic tools further constrained adherence. A provider explained,

> “When you consider the hospital’s setup, we have just one BP machine running around, and the monitor is faulty…. tests for renal failure aren’t done. The machine is there, but there’s an issue. They’re not being conducted yet; I think the lab isn’t fully operational” (Medical Doctor).

Reliance on external facilities for radiological investigations was also reported. A provider noted,

> “Regarding fundoscopy, we don’t do it here. Often, for ECHO and ECG tests, we have to send patients to the regional hospital, then they return with the results. Our hospital still doesn’t have the capacity to do these tests. The equipment and tools are there, but there are no specialists to conduct those tests, we are still negotiating, waiting” (Specialist Physician).

Heavy patient loads and limited beds further constrained care,

> “Sometimes there’s heavy congestion that impacts us. When a patient arrives, you might need to admit another, give drugs, and observe for some time to ensure the BP drops, then provide education before they leave. That’s tough when beds are scarce—we have only five, but the patient load is high” (Medical Doctor).

### Provider-Related Barriers

The participants reported limited formal training on the NSTGs in the management of Hypertension:

> “I think two or three doctors were taken for training in Dodoma. When they returned, they started that clinic for patients with diabetes and hypertension. But I’ve never seen if they came with a guideline” (Assistant Medical Officer).

Staff turnover reduced knowledge retention:

> “Of those doctors who went for training, only one has remained, the others were transferred. There, knowledge is lost” (Assistant Medical Officer).

Some providers relied more on experience than on consulting the NSTGs

> “Blood pressure medications are well-known; it’s almost routine. But to say I refer to the guideline when seeing a patient—rarely” (Medical Doctor).

### Patient-Related Barriers

Providers noted that patient financial limitations affected adherence.

> “For tests, those for patients with Hypertension, Diabetes Mellitus, these Non-Communicable diseases, are very costly… Fortunately, many common drugs are affordable. Nifedipine, Lasix, Spironolactone, Amlodipine aren’t expensive. The challenge arises when you want to move a patient from one drug group to an advanced, costlier group” (Medical Doctor).

Limited health literacy was also reported

> “You may track the patient, educate them, they understand, and you prescribe treatment. But on their next visit, you find they have complications. When you look at the medications you prescribed, they only got a few, and the others they couldn’t afford” (Clinical Officer).

Another provider highlighted knowledge gaps in the community

> “Many lack knowledge. There’s insufficient community education. For instance, a hypertensive patient, often of advanced age, doesn’t know their age is a Hypertension risk. When you explain or prescribe drugs, you can tell they haven’t accepted it; they might not take them at all” (Medical Doctor).

### Facilitators That Enhance Adherence Supportive Practices and Resources

Providers highlighted peer support, mentorship, and CME sessions as key facilitators.

> Structured CME sessions and mentorship reinforced adherence. “For training, we sometimes have doctors from programs like Mama Samia’s on-job training or from other facilities, which is good. We’ve learned a lot from Mama Samia’s program, where specialists come to mentor us. We ask questions, see where we’re going wrong, and improve… You can also learn on the job or visit other facilities to see how they treat patients and run their clinics” (Medical Doctor).

> Another provider added, “Yes, we have CME every Tuesday. We have different departments, and the presentations vary. For example, OPD might present on hypertension or malaria on a given Tuesday. First, it helps you not forget things. You might forget something, but the NSTGs helps in treating the patient properly, especially with dosage—someone might forget, or in an emergency, you can refer to it, and the patient gets the right treatment” (Assistant Medical Officer).

> On-the-job training was also valued: “Yes, I’ve had on job training. When you encounter a case and a senior doctor explains it to you, that’s on job training. If you see a similar patient the next day, you know where to start” (Clinical Officer).

> Digital access to the NSTGs in the management of Hypertension was noted as helpful: “I have a hardcopy, I was given one…but the online one is available, you can open GOTHOMIS and on the side open the STG because it’s there, if you search for it on the computer, it comes up. The challenges I see: first, availability of resources. Hard copies are scarce, so soft copies are needed since they’re quicker to access, especially with many patients” (Specialist Physician).

## Discussion

This study revealed that only 26.2% of hypertensive patients at public district hospitals in Dar es Salaam received management that was completely adherent to the NSTGs consistent with evidence from Dodoma where 29.29% of patients received guideline-concordant management across all conditions. (29) This reflects systemic challenges across public health facilities in Tanzania that hinder consistent application of standardized protocols.

Comparable trends have been observed in other sub-Saharan African settings, including Ethiopia (19.5%), Ghana (30%), and Zimbabwe (35%) (28,30,31), where inadequate provider training and resource limitations were key barriers to adherence. Conversely, higher adherence in better-resourced tertiary hospitals, such as 67.1% in Nigeria, underscores the critical role of institutional support, continuous professional development, and resource availability in facilitating evidence-based practice. (32)

Our findings indicate that patients attending follow-up visits were markedly more likely to receive guideline-concordant care, aligning with evidence from the United States and Indonesia that ongoing care enhances patients’ health literacy, thereby supporting consistent guideline application (33,34). Regular clinic visits likely allow providers to build familiarity with patients’ treatment plans, facilitating adherence, whereas initial consultations may challenge guideline application due to incomplete patient histories. Similarly, the presence of comorbidities roughly doubled the likelihood of guideline-concordant management, consistent with a finding in South Africa for Hypertensive patients with comorbid Diabetes Mellitus. (27) Furthermore, adherence was also higher in management of patients with Grade 1 and 2, compared to controlled patients although adherence for Grade 3 hypertension was not statistically significant, due to fewer number of patients. This shows that providers are more likely to adhere guidelines when faced with clinical complexities.

In this study healthcare provider adherence was not associated with patient age, sex, nor mode of payment. This contrasts with a study in the United States which suggested that demographic factors like age and gender may necessitate deviations from guidelines due to individualized needs (35).

Specialists showed a sevenfold higher prevalence of complete adherence compared to clinical officers, consistent with studies in Nigeria (32)and Malaysia(36). linking provider knowledge to guideline adherence (32,36). Their advanced training and exposure to evidence-based practices likely enable more accurate application of the NSTGs, whereas clinical officers, with less formal training, often rely on informal mentorship as qualitative data revealed. The wide confidence interval suggests limited precision, possibly due to small subgroup sizes, highlighting the need for larger studies to confirm this effect.

Providers with five or more years of experience showed 3 to 4-fold higher adherence compared to those with less than five years, consistent with a study in Sudan linking adherence to clinical experience (37), but contrasting a study in United States where less experienced providers relied heavily on guidelines (35). In Dar es Salaam, experienced providers likely develop practical skills enabling better NSTG application, while newer providers face challenges due to limited training.

Guideline-related factors emerged as significant barriers. Providers reported that the current edition of NSTGs is outdated, lacking updated drug regimens and protocols, and provided limited guidance for managing comorbidities, compelling reliance on clinical judgment or external references. The limited involvement of frontline providers in guideline development further reduced relevance to local practice, echoing findings from Zimbabwe that inclusive guideline development enhances adherence (38). Resource constraints, including persistent drug shortages, malfunctioning or unreliable blood pressure monitors, and frequent stock-outs of essential supplies, substantially hindered consistent application of the NSTGs. These challenges were compounded by high patient volumes and heavy workloads, which limited the time healthcare providers could dedicate to thorough clinical assessment and documentation. Inadequate supervision and irregular clinical audits further weakened accountability and opportunities for feedback, leading to variability in practice and reduced motivation to adhere strictly to guidelines.

Such systemic and structural barriers mirror broader evidence from sub-Saharan Africa, where the absence of supportive health system environments characterized by adequate staffing, continuous supervision, and reliable resource availability has been shown to undermine adherence to standard treatment guidelines and overall quality of care. (37,39–42).

Socioeconomic factors, including high out-of-pocket costs and low health literacy, also contributed to deviations from recommended care, underscoring the influence of patient context on provider adherence. Similar observations were reported in Zimbabwe, where financial barriers and limited awareness hindered blood pressure control (38), and in Nigeria, where inadequate insurance coverage and poor understanding of hypertension reduced quality of care (39). Studies from Ghana and Tanzania further highlight that limited patient knowledge and poor facility preparedness continue to impede effective hypertension management (40,43).

Facilitators of adherence included peer support and mentorship, access to digital NSTGs, and CME sessions, which enhanced guideline use even in resource-limited settings. Providers highlighted that peer consultation and informal mentorship helped in complex cases, while digital copies allowed rapid reference when hard copies were unavailable. This was similarly observed in Saudi Arabia and Malaysia (44,45).

Overall, these findings indicate that adherence to hypertension management guidelines in Dar es Salaam is shaped by patient complexity, provider expertise, guideline relevance, and systemic factors, including resources, supervision, and workload.

### Strengths and Limitations of the study

This study has several strengths. Its mixed-methods design, combining analysis of 397 patient files with 11 in-depth interviews, provides a comprehensive assessment of guideline adherence. Data collection was rigorous, using a structured checklist based on the 6th edition NSTGs and cross-verification with GOTHOMIS and manual registers, enhancing reliability. Conducted across five urban district hospitals, the study offers insights relevant to urban hypertension management. The integration of quantitative and qualitative data further strengthens the credibility and depth of the findings.

However, there are limitations. The cross-sectional design prevents causal inference, limiting understanding of temporal relationships between adherence and associated factors. Focusing solely on urban hospitals restricts generalizability to rural settings, where staffing, infrastructure, and patient characteristics may differ. Retrospective review of patient files introduced potential inaccuracies, with 16.2% of files excluded due to missing information. Additionally, non-pharmacological interventions, such as dietary counseling and lifestyle advice, were often under-documented, limiting the scope of assessment of healthcare provider adherence. The qualitative component did not differentiate responses by provider cadre, limiting identification of cadre-specific barriers, and patient perspectives were not included, preventing triangulation of provider reports. Although confidentiality and private interviews mitigated potential Hawthorne effects, self-reporting may have introduced bias, with providers possibly overstating adherence or underreporting challenges. Small subgroup sizes, particularly among patients attended by specialists, resulted in wide confidence intervals, reducing precision.

## Conclusion and Recommendations

Adherence to the National Standard Treatment Guidelines for hypertension management in public district hospitals in Tanzania remains suboptimal, increasing the risk of preventable complications. Several key barriers and facilitators influencing adherence were identified.

To improve practice, the Ministry of Health should regularly update the NSTGs every 2–3 years to reflect current evidence and local contexts, with active involvement of healthcare providers in guideline development, dissemination, and sensitization. Incorporating adherence indicators into supportive supervision and district performance assessments would strengthen accountability. In the longer term, expanding health insurance coverage and introducing subsidy mechanisms for essential diagnostics, medicines, and follow-up services could reduce out-of-pocket costs for patients.

Health facilities should enhance training, infrastructure, supply chains, access to guidelines, and supportive supervision, alongside routine audits. Further research is needed to assess adherence across diverse settings, include patient perspectives, and link provider practices to clinical outcomes. Coordinated policy, facility, and research efforts are essential to improve guideline adherence and hypertension outcomes nationwide.

## Data Availability

Due to ethical restrictions related to patient confidentiality and protection of sensitive personal health information, the datasets generated and/or analyzed during the current study are not publicly available. De-identified quantitative data and relevant excerpts from qualitative interviews are available from the corresponding author upon reasonable request and with permission from the relevant ethics review committee.

## Acknowledgements

The authors express their sincere gratitude to all who contributed to the success of this study. We extend our appreciation to the public district hospitals and district municipality authorities for their support and cooperation. We are deeply grateful to all study participants for their invaluable contributions, which were essential to the completion of this research. This study was conducted as part of a Master’s degree in Project Management, Monitoring, and Evaluation in Health at Muhimbili University of Health and Allied Sciences.

## Author Contributions

- Conceptualization: Gasper Singfrid Mung’ong’o
- Data curation: Gasper Singfrid Mung’ong’o, Paul Alikado Sabuni
- Formal analysis: Gasper Singfrid Mung’ong’o, Paul Alikado Sabuni, Gladys Mahiti
- Funding acquisition: Gasper Singfrid Mung’ong’o
- Investigation: Gasper Singfrid Mung’ong’o
- Methodology: Gasper Singfrid Mung’ong’o, Rose Mpembeni, Humphrey Godwin Medarakini
- Project administration: Gasper Singfrid Mung’ong’o
- Supervision: Rose Mpembeni, Daniel Msilanga
- Validation: Rose Mpembeni, Daniel Msilanga, Gladys Mahiti, Humphrey Godwin Medarakini
- Writing – original draft: Gasper Singfrid Mung’ong’o
- Writing – review & editing: Gasper Singfrid Mung’ong’o, Rose Mpembeni, Daniel Msilanga, Gladys Mahiti

## Notes

### Competing Interest Statement

The authors have declared no competing interest.

### Funding Statement

This research received no specific grant from any funding agency in the public, commercial, or not-for-profit sectors. The study was self-funded by the author as part of academic dissertation requirements.

### Author Declarations

MUHIMBILI UNIVERSITY OF HEALTH AND ALLIED SCIENCES (MUHAS) IRB

## References

1. World Health Organization. Global Health Estimates 2019: Deaths by cause, age, sex, by country and by region,2000–2019, Geneva, [Internet]. 2020 [cited 2024 Mar 10]. Available from: https://www.who.int/data/global-health-estimates

2. Zhou B, Carrillo-Larco RM, Danaei G, Riley LM, Paciorek CJ, Stevens GA, et al. Worldwide trends in hypertension prevalence and progress in treatment and control from 1990 to 2019: a pooled analysis of 1201 population-representative studies with 104 million participants. Lancet. 2021;398(10304):957–80.

3. Galson SW, Stanifer JW, Hertz JT, Temu G, Thielman NM, Gafaar T, et al. The Burden of Hypertension in the Emergency Department and Linkage to Care: A Prospective Cohort Study in Tanzania. PLoS One. 2019;14(1):e0211287.

4. Ivy A, Nguyen-Van-Tam JS, Dewhurst M, Gray WK, Chaote P, Rogathi J, et al. Ambulatory Blood Pressure Monitoring to Assess the White-Coat Effect in an Elderly East African Population. J Clin Hypertens. 2015;17(5):389–94.

5. Jones R, Putnam HWI, Philippin H, Cleland CM, Steel D, Gray WK, et al. Retinal Imaging to Identify Target Organ Damage in Older Africans: A Pilot Study. J Clin Hypertens. 2018;20(9):1296–301.

6. Lutambi AM. Prevalence and Determinants of Hypertension Among Adults of Reproductive Age in Tanzania: Analysis of a Cross-Sectional Demographic and Health Survey. BMJ Open. 2025;15(6):e094387.

7. Okello S, Muhihi A, Mohamed SF, Ameh S, Ochimana C, Oluwasanu AO, et al. Hypertension Prevalence, Awareness, Treatment, and Control and Predicted 10-Year CVD Risk: A Cross-Sectional Study of Seven Communities in East and West Africa (SevenCEWA). BMC Public Health. 2020;20(1).

8. Organization WH. Global report on hypertension: the race against a silent killer. World Health Organization; 2023.

9. Vos T, Lim SS, Abbafati C, Abbas KM, Abbasi M, Abbasifard M, et al. Global burden of 369 diseases and injuries in 204 countries and territories, 1990–2019: a systematic analysis for the Global Burden of Disease Study 2019. Lancet. 2020;396(10258):1204–22.

10. Assembly UNG. Transforming our world: the 2030 Agenda for Sustainable Development. 2015;

11. Cureau FV, Fuchs FD, Fuchs SCPC, Moreira LB, Schaan BD, Cisneros JZ, et al. Worldwide trends in hypertension prevalence and progress in treatment and control from 1990 to 2019: a pooled analysis of 1201 population-representative studies with 104 million participants. Lancet London Vol 398, no 10304 (Sep 2021), p 957-980. 2021;

12. Abeasi DA, Abugri D, Akumiah PO. Predictors of medication adherence among adults with hypertension in Ghana. J Client-Centered Nurs Care. 2022;8(1):23–32.

13. Ploth DW, Mbwambo J, Fonner VA, Horowitz BZ, Zager P, Schrader R, et al. Prevalence of CKD, Diabetes, And Hypertension in Rural Tanzania. Kidney Int Reports. 2018;3(4):905–15.

14. Muhihi AJ, Anaeli A, Mpembeni RNM, Sunguya BF, Leyna G, Kakoko D, et al. Prevalence, Awareness, Treatment, and Control of Hypertension among Young and Middle-Aged Adults: Results from a Community-Based Survey in Rural Tanzania. Morello F, editor. Int J Hypertens [Internet]. 2020;2020:9032476. Available from: 10.1155/2020/9032476

15. Trifirò S, Cavallin F, Mangi S, Mhaluka L, Maffoni S, Taddei S, et al. Hypertension in People Living With HIV on Combined Antiretroviral Therapy in Rural Tanzania. Afr Health Sci. 2023;23(1):129–36.

16. Mosha N, Mahande MJ, Juma A, Mboya IB, Peck RN, Urassa M, et al. Prevalence,awareness and Factors Associated With Hypertension in North West Tanzania. Glob Health Action. 2017;10(1).

17. Aytenew TM, Kassaw A, Simegn A, Nibret Mihretie G, Asnakew S, Tesfahun Kassie Y, et al. Uncontrolled hypertension among hypertensive patients in Sub-Saharan Africa: A systematic review and meta-analysis. PLoS One. 2024;19(6):e0301547.

18. Dewhurst MJ, Dewhurst F, Gray WK, Chaote P, Orega GP, Walker RW. The high prevalence of hypertension in rural-dwelling Tanzanian older adults and the disparity between detection, treatment and control: a rule of sixths? J Hum Hypertens. 2013;27(6):374–80.

19. Maginga J, Guerrero M, Koh E, Holm Hansen C, Shedafa R, Kalokola F, et al. Hypertension control and its correlates among adults attending a hypertension clinic in Tanzania. J Clin Hypertens. 2016;18(3):207–16.

20. Mwita JM, Nyawale HA, Mghanga FP. The burden of hypertension and its associated factors among adults in Ruvuma, Southern Tanzania. 2020;

21. Republic U. United Republic of Tanzania National Strategic Plan for Prevention and Control of Non-Communicable Diseases 2021-2026 National Strategic Plan for Non-Communicable Diseases. 2026;

22. Urassa DP, Mwangu M, Makwi JK. Utilization of standard treatment guidelines (STG) at primary health facilities, Magu district, Tanzania. East Afr J Public Health. 2013;10(1):238–45.

23. National Medicine and Therapeutic Committee. Standard Treatment Guidelines Tanzania. 2021;Sixth edit:596. Available from: https://medicine.st-andrews.ac.uk/igh/wp-content/uploads/sites/44/2022/01/STG-NEMLIT-2021.pdf

24. Garg R. Individualization of Hypertension Treatment: An Expert Review. Int J Adv Med. 2023;10(8):647–52.

25. Long AN, Dagogo-Jack S. Comorbidities of Diabetes and Hypertension: Mechanisms and Approach to Target Organ Protection. J Clin Hypertens. 2011;13(4):244–51.

26. ministry of health tanzania. Hospitals at district level [Internet]. 2024 [cited 2024 Nov 9]. Available from: https://hfrs.moh.go.tz/web/index.php?r=portal%2Fquick-search&filters=HOSD

27. Adedeji AR, Tumbo J, Govender I. Adherence of doctors to a clinical guideline for hypertension in Bojanala district, North-West Province, South Africa. African J Prim Heal Care Fam Med. 2015;7(1):1–6.

28. Ataro BA, Mulatu G, Mengistu D. Compliance With Guidelines of Hypertension Management, and Associated Factors Among the Health Practitioners. Inq J Heal Care Organ Provision, Financ. 2023;60:00469580231216400.

29. Wiedenmayer K, Ombaka E, Kabudi B, Canavan R, Rajkumar S, Chilunda F, et al. Adherence to standard treatment guidelines among prescribers in primary healthcare facilities in the Dodoma region of Tanzania. BMC Health Serv Res. 2021;21:1–10.

30. Basopo V, Mujasi PN. To what extent do prescribing practices for hypertension in the private sector in Zimbabwe follow the national treatment guidelines? An analysis of insurance medical claims. J Pharm Policy Pract. 2017;10(1):37.

31. Harrison MA, Marfo AFA, Buabeng KO, Annan A, Nelson F, Boateng DP, et al. Blood Pressure—lowering Medication Prescribing, Its Adherence to Guidelines and Relationship With Blood Pressure Control at a Family Medicine Department. Heal Sci Reports. 2023;6(4).

32. Ibezim IC, Naylor Ian, Isah A, Igboeli Nu. Adherence to Essential Hypertension Treatment Guidelines in a Tertiary Hospital in Nigeria. Int J Pharm Pharm Sci. 2020;106–10.

33. Lu Y. Barriers to Optimal Clinician Guideline Adherence in the Management of Markedly Elevated Blood Pressure: A Qualitative Content Analysis of Electronic Health Records. 2024;

34. Mulyanto J, Kringos D, Kunst AE. Socioeconomic Inequalities in the Utilisation of Hypertension and Type 2 Diabetes Management Services in Indonesia. Trop Med Int Heal. 2019;24(11):1301–10.

35. Cabezas S, Grant K, Clark J. Evaluating the Prescribing Patterns of Antihypertensive Medications in a Student-Run Free Clinic. J Student-Run Clin. 2023;9(1).

36. Ahmad N, Khan AH, Khan I, Khan A, Atif M. Doctors’ knowledge of hypertension guidelines recommendations reflected in their practice. Int J Hypertens. 2018;2018(1):8524063.

37. Abdelgadir HS, Elfadul MM, Hamid NH, Noma M. Adherence of doctors to hypertension clinical guidelines in academy charity teaching hospital, Khartoum, Sudan. BMC Health Serv Res. 2019;19:1–6.

38. Goverwa TP, Masuka N, Tshimanga M, Gombe NT, Takundwa L, Bangure D, et al. Uncontrolled hypertension among hypertensive patients on treatment in Lupane District, Zimbabwe, 2012. BMC Res Notes. 2014;7:1–8.

39. Odusola AO, Stronks K, Hendriks ME, Schultsz C, Akande T, Osibogun A, et al. Enablers and barriers for implementing high-quality hypertension care in a rural primary care setting in Nigeria: perspectives of primary care staff and health insurance managers. Glob Health Action. 2016;9(1):29041.

40. Boima V. Level of Understanding and Community-Level Barriers to the Management of Hypertension: A Qualitative Study in Eight Coastal Communities in Ghana. BMJ Glob Heal. 2025;10(8):e017511.

41. Commodore–Mensah Y, Liu X, Ogungbe O, Ibe CA, Amihere J, Mensa M, et al. Design and Rationale of the Home Blood Pressure Telemonitoring Linked With Community Health Workers to Improve Blood Pressure (LINKED-BP) Program. Am J Hypertens. 2023;36(5):273–82.

42. Philbert R, Temba PM, Ebrahim AA, Rusobya H, Mashili F. Hypertension Beyond the Clinic: Patient Perspectives on Self-Management in a Resource-Limited Urban Setting. 2025;

43. Adinan J, Manongi R, Temu GA, Kapologwe N, Marandu A, Wajanga B, et al. Preparedness of health facilities in managing hypertension & diabetes mellitus in Kilimanjaro, Tanzania: a cross sectional study. BMC Health Serv Res. 2019;19:1–9.

44. Shnaimer JA, Gosadi IM. Primary health care physicians’ knowledge and adherence regarding hypertension management guidelines in southwest of Saudi Arabia. Medicine (Baltimore). 2020;99(17):e19873.

45. Ahmad N, Tangiisuran B, Meng OL, Aziz NA, Ahmad F, Atif M. Guidelines Adherence and Hypertension Control at a Tertiary Hospital in Malaysia. J Eval Clin Pract. 2012;19(5):798–804.

